# Non contrast-enhanced imaging as a replacement for contrast-enhanced imaging for MRI automatic delineation of nasopharyngeal carcinoma

**DOI:** 10.1101/2020.07.09.20148817

**Authors:** Lun M. Wong, Qi-yong H. Ai, Frankie K.F. Mo, Darren M.C. Poon, Ann D. King

**Affiliations:** Department of Imaging and Interventional Radiology, The Chinese University of Hong Kong, Prince of Wales Hospital, Hong Kong SAR; Department of Clinical Oncology, State Key Laboratory of Translational Oncology, The Chinese University of Hong Kong, Hong Kong SAR

## Abstract

**Purpose:** Convolutional neural networks (CNNs) show potential for delineating cancers on contrast-enhanced MRI. However, there is world-wide interest in reducing the administration of MRI contrast agents. We aim to determine if CNNs can automatically delineate primary nasopharyngeal carcinoma (NPC) using the non contrast-enhanced (NE) T2-weighted fat-suppressed (T2W-FS) sequence and compare the performance with that from the contrast-enhanced T1-weighted (CE-T1W) sequence.

**Materials and methods:** We retrospectively analyzed primary tumors in 201 patients with NPC. Six patients were randomly sampled as the training-validation group to avoid over-fitting, and the remaining 195 patients underwent 3-fold cross-validation analysis. We trained and tested a well-established two-dimensional CNN, U-Net, for tumor delineation on CE-T1W and T2W-FS sequences. CNN-derived delineations on CE-T1W and T2W-FS were compared to manual delineation using the Dice similarity coefficient (DSC) and average surface distance (ASD). Differences in DSC and ASD of CNN-derived delineations between CE-T1W and T2W-FS sequences were compared using the Wilcoxon rank test.

**Results:** The CNN’s tumor delineation performance on CE-T1W and T2W-FS were similar in DSC (0.71±.09 vs. 0.71±.09, *p=*0.50) and ASD (0.21±0.48cm vs. 0.17±0.19cm, *p=*0.34).

**Conclusion:** CNN can automatically delineate primary NPC using the NE T2W-FS sequence, which has the potential to substitute the CE-T1W sequence.

## Introduction

Primary tumor delineation on magnetic resonance imaging (MRI) is an essential step for cancer staging and treatment planning [1–6]. More recently, it has also become important for quantitative analysis that aids the prediction and monitoring of treatment response [7–12]. Regardless of its aim, primary tumor delineation is a laborious and demanding task to perform manually.

Convolutional neural networks (CNNs) have shown promise for the MRI-based delineation of malignant tumors in the brain, lung and pancreas [13–16]. Nasopharyngeal carcinoma (NPC) is a particularly challenging cancer to delineate because its boundaries can have complex anatomy owing to the many different types of tissues in the surrounding region, including the bone of the skull base. Previous literature has reported successful CNN adaptations in the automatic delineation for primary NPC [17–21], but the work to-date has relied on gadolinium-based contrast-enhanced (CE) MRI to optimize the result. In addition to the extra scanning time and monetary cost, gadolinium-based contrast agents are being used more sparingly now that gadolinium is known to deposit in the human body, including the brain [22], and the long term effects of this deposition are unknown. Therefore, a non contrast-enhanced (NE) substitution of CE sequences for primary tumor delineation is desirable. It would be especially advantageous in patients who undergo multiple MRI examinations or have impaired renal function, as well as for monitoring intra-treatment response.

The T2-weighted fat-suppressed (T2W-FS) is a promising NE substitute to the contrast-enhanced T1-weighted (CE-T1W) MRI for primary tumor delineation. The T2W-FS sequence is not only a widely available, well-established sequence and part of the routine NPC protocol, but is also effective at depicting tumor boundaries against many different types of normal tissues. These tissues include the bony skull base when T2-weighted imaging is combined with the suppression of fat signal in the bone marrow. Furthermore, previous literature has highlighted the diagnostic values of the T2W-FS in evaluating soft-tissue tumor extent [23,24] and cancer staging [25].

Therefore, the purpose of this study is to provide a comparison between NPC primary tumor delineation performance of CNN on CE-T1W and T2W-FS images, with the performance evaluated with reference to manual delineation by an expert on NPC, and to determine whether T2W-FS images can serve as a substitute to CE-T1W images for automatic primary tumor delineation.

## Materials and methods

### Patients

This retrospective study was approved by the local institutional board, and the requirement of written consent was waived. During 2010 to 2013, 201 patients (age: 54.5 ± 11.5 years; 157 men and 44 women), with newly diagnosed biopsy-proven undifferentiated NPC who underwent head and neck MRI for staging and were scanned with axial T2W-FS and CE-T1W, were included for the analysis. The patients with NPC were staged T1, T2, T3, and T4 in 67, 29, 73, and 32 patients respectively based on the 8^th^ edition of the AJCC Cancer Staging Manual [3].

### Imaging acquisition

MRI was performed using a Philips Achieva TX 3T scanner (Philips Healthcare, Amsterdam, Netherland). The protocol included (a) an axial fat-suppressed T2-weigthed turbo spin-echo sequence (repetition time /echo time, 4000/80 msec; field of view, 230 × 230 mm; section thickness, 4 mm; echo train length, 15-17; sensitivity encoding factor, 1; number of signal acquired, 2) and (b) an axial T1-weighted turbo spin-echo sequence (repetition time /echo time, 500/10 msec; field of view, 230 × 230 mm; section thickness, 4 mm; echo train length, 4; sensitivity encoding factor, 1; number of signal acquired, 2) following a bolus injection of 0.1 mmol of gadoteric acid (Dotarem; Guerbet, Villepinte, France) per kilogram of body weight.

### Manual delineation of primary NPC

All primary NPC tumors were manually delineated on the axial CE-T1W and T2W-FS images with references to all series of pre- and post-contrast MRI sequences available. Manual delineation was performed by a researcher with 6 years of experience in MRI of NPC using the opensource software ITK-SNAP v3.4.0 [26]. Manual delineation was necessary on both sequences as minor patient inter-scans movements can translate to a substantial displacement of tumor outline, especially for small early-NPCs. Dice similarity coefficient (DSC) and average surface distance (ASD) were used to show the differences between manually drawn labels on the two sequences. The primary tumor volume (PTV) was calculated by multiplying the voxel size with the number of voxels labelled as primary NPC.

### Comparing delineation performance of CNN between the two sequences

Of the 201 cases, 6 were randomly sampled as the training-validation set, which refers to a small set of data unseen during training to monitor whether over-fitting occurred as described in [27], and the remaining 195 cases were analyzed with 3-fold cross-validation. In each fold, the designated CNN architecture, U-Net [28], was trained from scratch with 130 training cases and then tested on 65 cases twice, once with CE-T1W images and once with T2W-FS images, with identical training parameters, as detailed in Table 1. In each epoch, each slice of an image volume were augmented into three additional slices by random rotations, scaling and gaussian-noise for improved training quality. The networks were implemented and trained by minimizing weighted cross-entropy loss with the stochastic gradient descent technique using the opensource Python package PyTorch v1.4 [29].

**Table 1.**
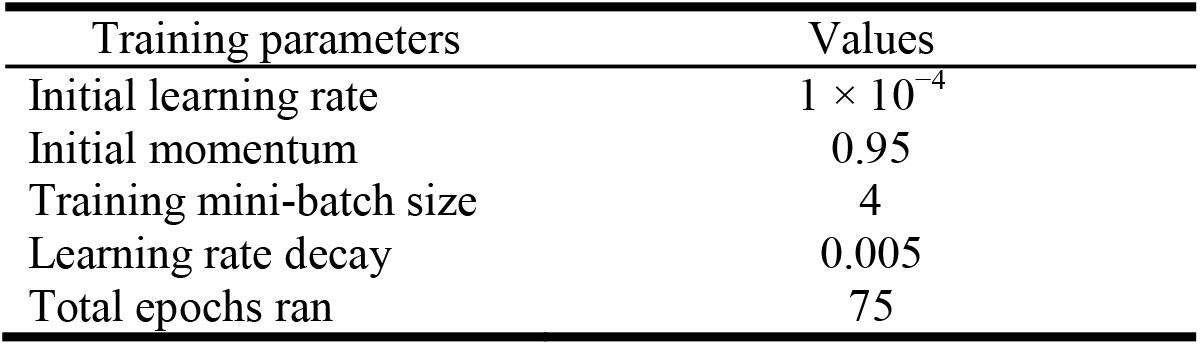
Key training parameters

The performance of CNN-based automatic delineation on CE-T1W and T2W-FS images were evaluated using the DSC and ASD, computed with respect to the manual delineation delineated by the expert. The PTVs from both the CNN-derived and manual delineations on each sequence were calculated by multiplying the voxel counts labeled as tumor to the voxel size. Details of these performance metrics can be found in [30].

The flow of the experiment is illustrated in Figure 1.

**Figure 1.**
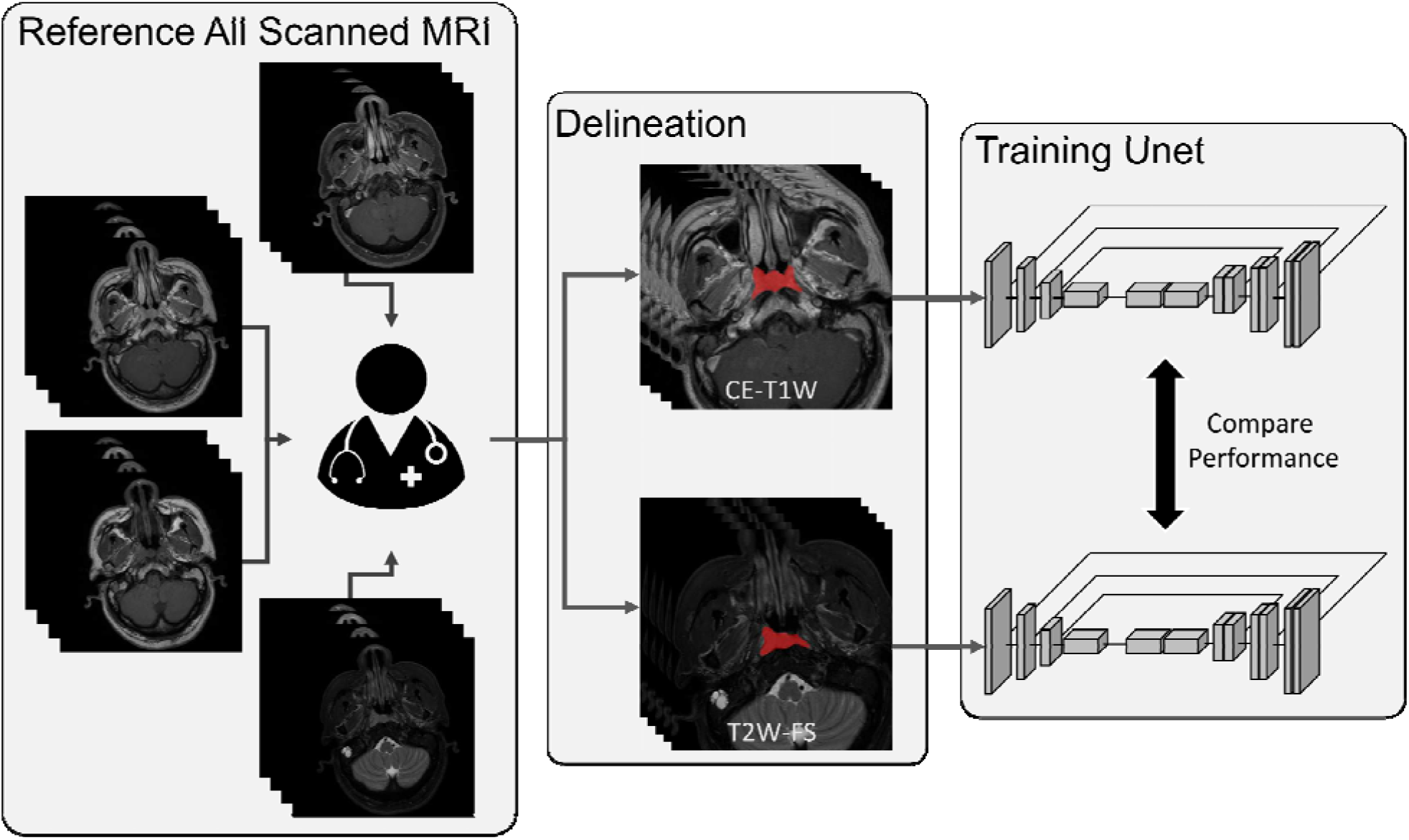
Flow chart of the experimental setup. Our expert delineated the primary tumor of NPC on both CE-T1W and T2W-FS images with reference to all available series of pre- and post-contrast MRI scans. U-Net was trained twice separately from scratch to delineate primary NPC, once on the CE-T1W and once on the T2W-FS images. The performance of the network wa computed with reference to the expert’s delineation and then compared between the sequences. We hypothesized that both CE-T1W and T2W-FS contain adequate information for the delineation of primary NPC such that image features on T2W-FS, though occult to human, can be detected by the U-Net. Deviation from this hypothesis would be reflected by the difference in delineation performance between the two trained networks. (NPC = nasopharyngeal carcinoma; CE-T1W = contrast-enhanced T1-weighted; T2W-FS = T2-weighted fat-suppressed)

### Statistical analysis

To verify the consistency of the manually delineated contours across sequences, the PTVs of the two sets of manual delineation were compared and their agreement was evaluated using the paired t-test and intra-class correlation (ICC).

The non-parametric one-way analysis of variance on ranks (Kruskal-Wallis H test) was performed to confirm the inter-fold consistency of DSC and ASD across the three folds on CE-T1W and T2W-FS images.

Differences in CNN’s tumor delineation performance between T2W-FS and CE-T1W were evaluated with the Wilcoxon rank test and Bland-Altman analysis [31]. In addition, differences in the PTVs between the CNN-derived and manual delineations on each sequence were also compared using the Wilcoxon rank test.

All statistical analysis were performed using IBM SPSS Statistics for Windows, version 25.0 (IBM, Armonk, USA) and open-source R library package blandr [32]. The significance level of differences was accepted at *p* < 0.05.

## Results

### Consistency of manual delineation across sequences

Regarding manual delineation, the difference in the PTVs between CE-T1W and T2W-FS was not significant (23.5 ± 26.7 cm^3^ and 23.1 ± 26.1 cm^3^ respectively, n = 201, *p =* 0.06) with an ICC of 0.996 (*p* < 0.05). The manual delineation on CE-T1W and T2W-FS had a DSC of 0.83 and ASD of 0.06 cm when compared against each other.

### Consistency of CNN performance across folds

The results of CNN’s tumor delineation performance across folds are tabulated in Table 2. No differences were observed in the DSC and ASD across folds on the CE-T1W (*p* = 0.84 and 0.78 respectively, n = 65 in each fold) and T2W-FS (*p* = 0.56 and *p* = 0.50 respectively) sequence.

**Table 2.** **Non-parametric** One-way ANOVA on ranks (Kruskal-Wallis H test) of CNN performance metrics across folds.

| Performance Metric | Means (n=65) |  |  | ANOVA<br>*p*-value |
| --- | --- | --- | --- | --- |
|  | Fold 1 | Fold 2 | Fold 3 |  |
| CE-T1W |  |  |  |  |
| DSC | 0.70 ± 0.09 | 0.71 ± 0.09 | 0.71 ± 0.10 | 0.84 |
| ASD (cm) | 0.18 ± 0.17 | 0.25 ± 0.77 | 0.21 ± 0.27 | 0.78 |
| T2W-FS |  |  |  |  |
| DSC | 0.71 ± 0.09 | 0.70 ± 0.08 | 0.70 ± 0.11 | 0.56 |
| ASD (cm) | 0.19 ± 0.21 | 0.15 ± 0.13 | 0.17 ± 0.21 | 0.50 |

Data are presented as mean ± standard deviation. ANOVA = analysis of variance, ASD = average surface distance, DSC = Dice similarity score, CNN = convolutional neural network, CE-T1W = contrast-enhanced T1-weighted, T2W-FS = T2-weighted fat-suppressed.

### Comparison of CNN performance difference across sequences

The performance metrics of CNN’s tumor delineation performance for CE-T1W and T2W-FS are tabulated in Table 3 and illustrated as boxplots in Figure 2, the corresponding Bland-Altman plot is provided in Figure 3. No significant differences were observed in the DSC and ASD between CE-T1W and T2W-FS (*p* = 0.50 and 0.34 respectively) (Table 3). The PTVs obtained from the CNN-based delineation were significantly larger on CE-T1W than that on T2W-FS (26.3 ± 25.5 cm^3^ and 24.2 ± 23.7 cm^3^ respectively, *p* < 0.001).

**Table 3.** Wilcoxon rank-test and Bland-Altman analysis of CNN performance across sequences. The paired t-test shows there are significant bias for the PTVs across sequences but not for the DSC and ASD.

| Performance Metric<br>(n = 195) | CE-T1W | T2W-FS | Bias | 95% Confidence Interval |  | *p*-value |
| --- | --- | --- | --- | --- | --- | --- |
|  |  |  |  | Lower Bound | Upper Bound |  |
| DSC | 0.71 ± 0.09 | 0.71 ± 0.09 | 0.001 | -0.01 | 0.02 | 0.50 |
| ASD (cm) | 0.21 ± 0.48 | 0.17 ± 0.19 | 0.04 | -0.03 | 0.11 | 0.34 |
| PTV (cm<sup>3</sup>) | 26.3 ± 25.5 | 24.2 ± 23.7 | 2.1 | 0.75 | 3.4 | **< 0.001** |

Data are presented as mean ± standard deviation. Bold face indicates statistical significance. ASD = average surface distance, DSC = Dice similarity score, CNN = convolutional neural network, CE-T1W = contrast-enhanced T1-weighted, T2W-FS = T2-weighted fat-suppressed, PTV = primary tumor volume.

**Figure 2.**
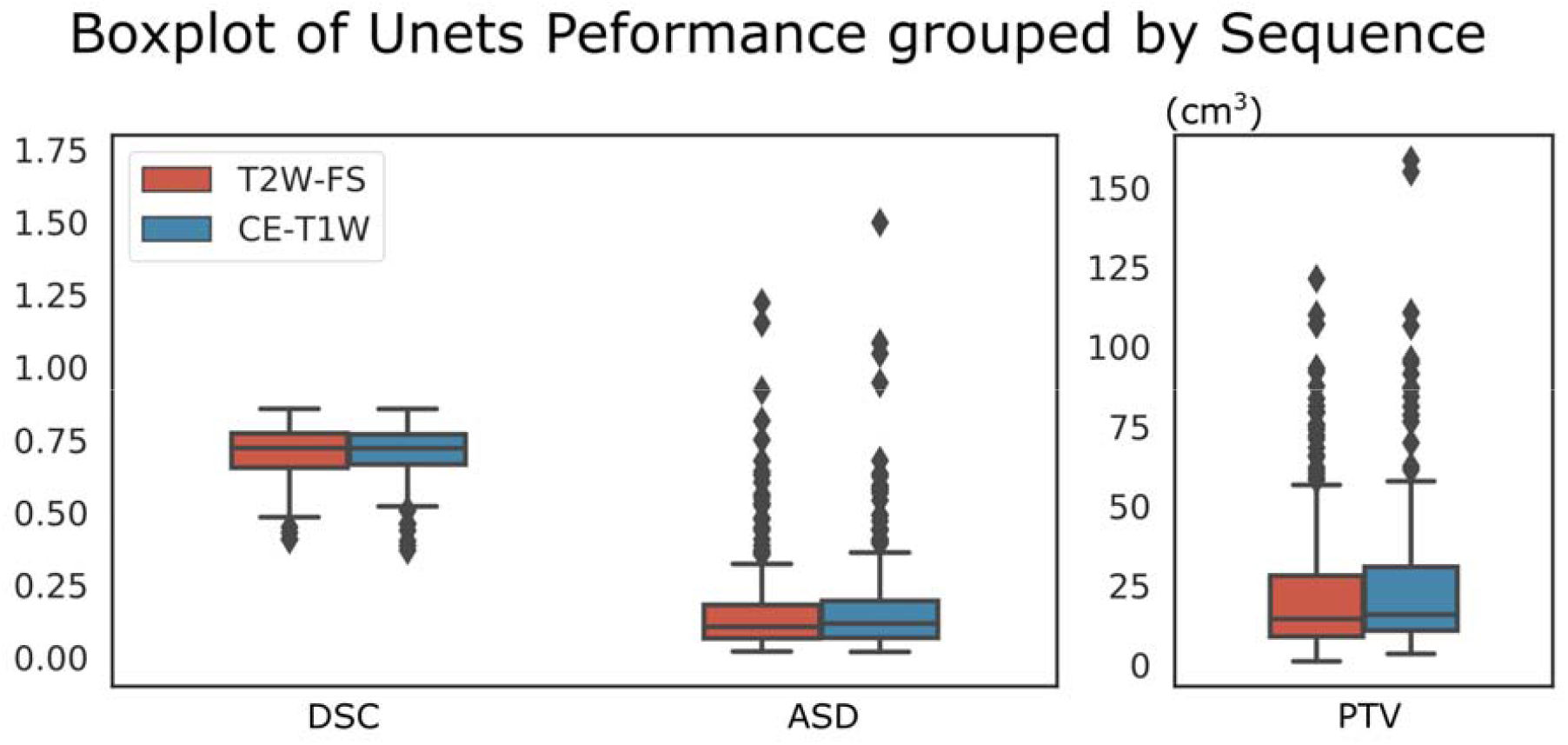
A boxplot showing the distribution of performance metrics (DSC, ASD and PTVs) of U-Nets for primary tumor delineation on CE-T1W and T2W-FS images. The plot highlights similarity in the distribution of performance when the CNN was trained to delineate the primary NPC on CE-T1W and T2W-FS images, however, the paired-sample t-test revealed a significant difference in the PTVs between sequences (26.3 ± 25.5cm^3^ vs. 24.2 ± 23.7cm^3^, respectively, *p* < 0.001) but not in the DSC (0.71 ± 0.09 vs. 0.71 ± 0.09 respectively, *p =* 0.50) and ASD (0.21 ± 0.48 cm vs. 0.17 ± 0.19 respectively, *p =* 0.34). (ASD = average surface distance; DSC = Dice similarity score; PTV = primary tumor volume; CE-T1W = contrast-enhanced T1-weighted; T2W-FS = T2-weigthed fat-suppressed)

**Figure 3.**
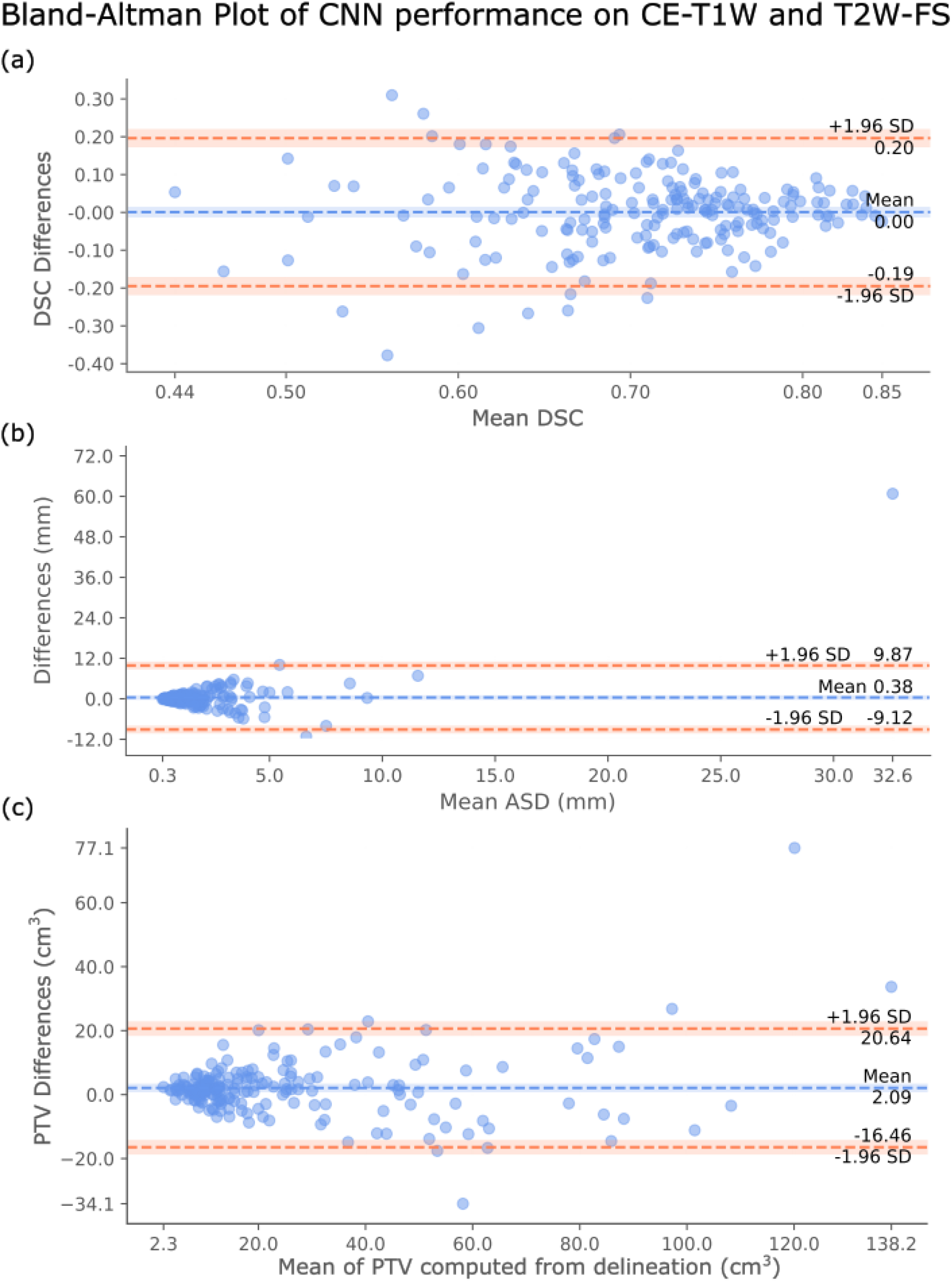
The Bland-Altman plot showing the agreements of CNN’s tumor delineation performance on CE-T1W and T2W-FS images in terms of the (a) DSC, (b) ASD and (c) PTV. The mean, upper (mean + 1.96 SD) and lower (mean − 1.96 SD) bounds of the differences in each plot are computed with 95% confidence interval shaded in corresponding colors. The exact values are labelled next to the indicating lines. The biases are close to zero in the three plots. A distinct point in (b) corresponds to a case where the U-Net have falsely labeled tumor tissues on a slice at the neck, far away from the tumor center on CE-T1W, resulting in large ASD. This sort of error was not seen on T2W-FS. (ASD = average surface distance; DSC = Dice similarity coefficient; PTV = primary tumor volumes; CE-T1W = contrast-enhanced T1-weighted; T2W-FS = T2-weighted fat-suppressed)

The PTVs obtained from CNN-derived delineation were significantly larger than those obtained from manual delineation on both CE-T1W (26.3 ± 25.5 cm^3^ and 23.5 ± 26.6 cm^3^ respectively, *p* < 0.001) and T2W-FS (24.2 ± 23.7 cm^3^ and 23.2 ± 26.2 cm^3^ respectively, *p <* 0.001). A representative example of both CNN-derived and manual delineations for primary NPC is shown in Figure 4.

**Figure 4.**
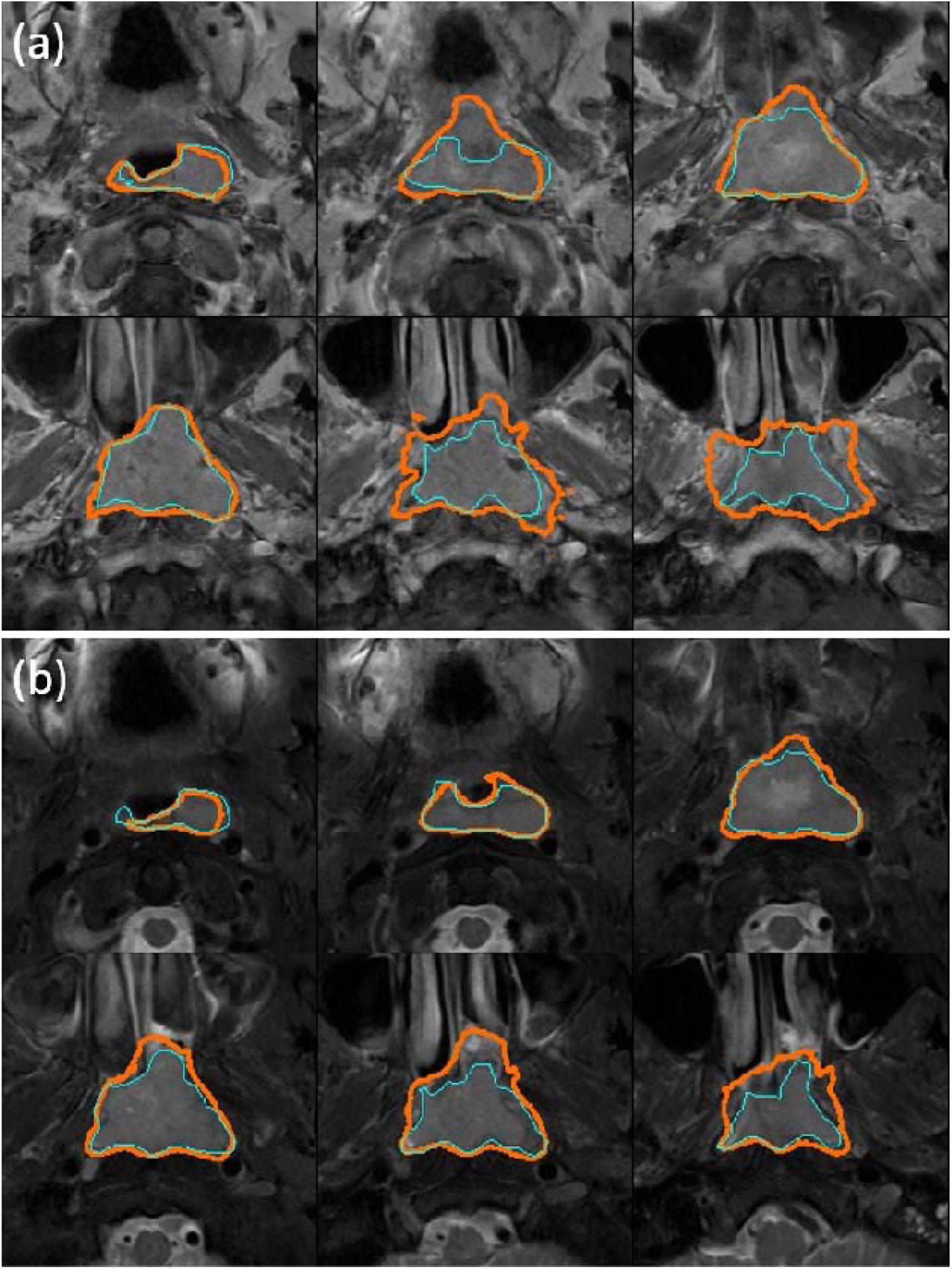
A representative example of the CNN-derived primary tumor delineation showing a case of NPC confined to the nasopharynx, trained and delineated on (a) the CE-T1W sequence, with a DSC of 0.68 and (b) the T2W-FS sequence, with a DSC of 0.69. The orange delineation was performed by the CNN and the light blue delineation was performed by an expert in th MRI of NPC. The DSC are similar for CE-T1W and T2W-FS sequences, but CNN-derived delineation was more prone to overestimation on the CE-T1W sequence than on T2-FS sequence. (CNN = convolutional neural network; DSC = Dice similarity coefficient; NPC = nasopharyngeal carcinomas; CE-T1W = contrast-enhanced T1-weighted; T2W-FS = T2-weigthed fat-suppressed)

## Discussion

This preliminary study investigated the performance of CNN-based automatic delineation of primary NPC on CE (CE-T1W) and NE (T2W-FS) MRI sequences. The results showed no significant difference in the performance of CNN-based tumor delineations between the two sequences. This indicates the NE T2W-FS images could be a potential substitute for CE-T1W images when using CNN for the automatic delineation of primary NPC. Most previous studies on CNN-based primary tumor delineation have relied on the CE MRI [17–19,33–35], but our results are encouraging and suggest that future adaptations of CNN for NE sequence are warranted to facilitate the reduction of contrast administration for MRI where possible.

With regard to the performance of the CNN primary NPC delineation using CE imaging, our results (DSC, 0.71; ASD, 2.1 mm), are better than those previously reported for U-Net and similar or slightly worse than those reported for using a customized CNN. We used U-Net as our testing reference because it is one of the most general and representative 2D delineation CNN architectures and its encoder-decoder design is the back-bone of many proposed delineation CNN architectures [15,18,34–38]. Only one other NPC study used the U-Net for primary tumor delineation and reported a DSC of 0.59 and ASD of 6 mm [18] on CE images. Using customized CNNs, three previous studies reported a mean/median DSC of 0.72-0.79 [17,18,20] and ASD of 2.0-2.1mm [17,18], while two studies, which included only 29 and 30 patients reported a higher DSC of 0.89 and 0.83 on CE imaging respectively [19,21]. Only one of these studies also tested their customized network on NE T2-weighted images, they reported a slightly lower DSC of 0.64 than our results and showed a dual-sequence input combining CE T1-weighted and NE T2-weighted sequences performed better than either sequences alone, but this study did not directly compare separate performance on the two sequences. Although U-Net showed similar performance metrics (in terms of DSC and ASD) in primary tumor delineation on CE-T1W and T2W-FS sequences in our study, it overestimated the PTV on both sequences and was more prone to overestimating the primary tumor extent on the CE-T1W than on the T2W-FS sequence. This could be explained by the elevated intensity profiles of CE images in general, which could have added to the probability of falsely including tissues surrounding the tumor, leading to the overestimation of the PTV. Unfortunately, the comparison of PTVs derived from CNN and manual delineation are rarely reported in the literature.

Our study has some limitations. First, although our results provide a valuable insight into CNNs and show that feature learning is not dependent on contrast enhancement, our U-Net results may not be generalizable to other CNN architectures. Second, we performed our test with a slice-based algorithm and did not consider other input configurations, such as patch-based or volumetric-based algorithms. Third, we did not verify our results with external data in this preliminary study.

In conclusion, the CNN, U-Net, adapted for primary NPC delineation in this study exhibited similar performance on CE-T1W and T2W-FS sequences and showed closer estimation of PTVs to those obtained from manual delineation on the T2W-FS sequence. The NE T2W-FS images can serve as a potential substitute to CE-T1W images for the purpose of automatic primary tumor delineation in patients with NPC.

## Data Availability

Raw data used in this study are under patient privacy protection and will not be available to readers, the derived-data will be provided to readers on demand.

